# Children’s Residential Proximity, Spousal Presence and Modifiable Risk Factors for Dementia among Older Adults with Cognitive Impairment

**DOI:** 10.1101/2023.10.24.23297470

**Authors:** Zhuoer Lin, Xuecheng Yin, Becca R. Levy, Yue Yuan, Xi Chen

## Abstract

**Background:** Cognitive impairment in older adults poses considerable challenges, and the role of family support becomes increasingly crucial. This study aims to examine the impact of children’s residential proximity and spousal presence on the key modifiable risk factors for dementia among older adults with cognitive impairment.

**Methods:** Utilizing the Health and Retirement Study (HRS) data from 1995 to 2018, we analyzed 14,731 participants (35,840 person-waves) aged 50 and older with cognitive impairment. Family support was characterized based on the presence of a spouse and residential proximity to children. Smoking, depressive symptoms and social isolation were included as the key modifiable risk factors for dementia identified in later life. Using mixed-effects logistic regressions, associations between access to family support and the modifiable risk factors were determined, adjusting for various socio-demographic and health-related factors.

**Results:** Significant associations were found between access to family support and modifiable risk factors for dementia. Cognitively impaired older adults with less available family support, characterized by distant-residing children and the absence of a spouse, had significantly higher risks of smoking, depressive symptoms, and social isolation. Moreover, we revealed a consistent gradient in the prevalence of the risk factors based on the degree of family support. Relative to older adults with a spouse and co-resident children, those without a spouse and with all children residing further than 10 miles displayed the highest risks of smoking, depressive symptoms, and social isolation.

**Conclusion:** Access to family support, particularly from spouses and proximate children, plays a protective role against key modifiable risk factors for dementia in older adults with cognitive impairment. The findings highlight the need for bolstering family and social support systems to enhance the well-being of this vulnerable population.

## 1. Introduction

Cognitive impairment, ranging from mild cognitive impairment to dementia, represents pressing challenges in the aging population, with far-reaching implications for individuals and their families.^1–3^ Older adults with cognitive impairment are particularly vulnerable as they are faced with extensive cognitive challenges while navigating a complex landscape of risk factors that are potentially modifiable.^1–6^ Their diminished cognitive capacity can heighten the difficulty of recognizing these risk factors, making informed decisions, and seeking appropriate social services and support.^1–6^

Amid the mounting number of persons with cognitive disorders, the role of family support becomes indispensable.^1–4,7^ Lack of such support has been associated with elevated risks of adverse outcomes, such as untreated medical conditions, self-neglect, malnutrition, and falls.^8–10^ As the bedrock of support systems, the family holds the potential to mitigate the risks associated with cognitive impairment and its associated risk factors. The presence of close family members, such as spouses and children, can act as a protective buffer, offering emotional, psychological, and practical support and assistance, especially for those with cognitive impairment.^4–7,11–14^ A nuanced comprehension of family support dynamics, encompassing the proximity of offspring and the presence of a spouse, thus becomes crucial in deciphering their influence on modifiable risk factors tied to dementia.^5,7,11,15^ However, there is no evidence examining the relationship between family support and modifiable risk factors for dementia among older adults living with cognitive impairment, particularly when considering the residential proximity to their children.

This study, utilizing longitudinal survey data of Americans aged 50 and above, delved into the proximity of children and spousal presence as indicators of family support and explored how they buffer against the development of modifiable risk factors within the context of older adults facing cognitive impairment. Specifically, the *Lancet* Commission Report on dementia prevention provided an established framework for approaching the research question.^1^ This systematic review synthesized existing literature and identified three leading modifiable risk factors in later life that wield the greatest influence over dementia (as measured by population attributable fraction for dementia): smoking, depressive symptoms, and social isolation.^1^ In this study, we focused on these three key modifiable risk factors, and hypothesized that cognitively impaired individuals with limited access to family support are at elevated risk of smoking, and experiencing depressive symptoms and social isolation.^1,16^

## 2. Methods

We analyzed data from the Health and Retirement Study (HRS), a nationally representative longitudinal survey of Americans aged 50 and older conducted biennially. For our study, we focused on participants surveyed between 1995 and 2018. This was because valid cognitive classifications were available from wave 1995 onwards;^17,18^ and 2018 was the last survey wave before the COVID-19 pandemic. We restricted the sample to community-dwelling older adults who were 50 years or older, had at least one living child, and exhibited cognitive impairment at the time of their interview. Cognitive impairment was assessed using a 27-point cognitive scale, and participants were categorized based on well-established criteria.^17,18^ This included those classified as “cognitively impaired but not demented” (scoring between 7-11 points) and those identified as “demented” (scoring between 0-6 points).^17,18^ We focused our analysis on participants who had complete data for covariates and modifiable risk factors. This resulted in a total sample of 14,731 persons and 35,840 person-waves (an average of 2.4 waves per person). The sample selection process is outlined in Figure 1.

**Figure 1.**
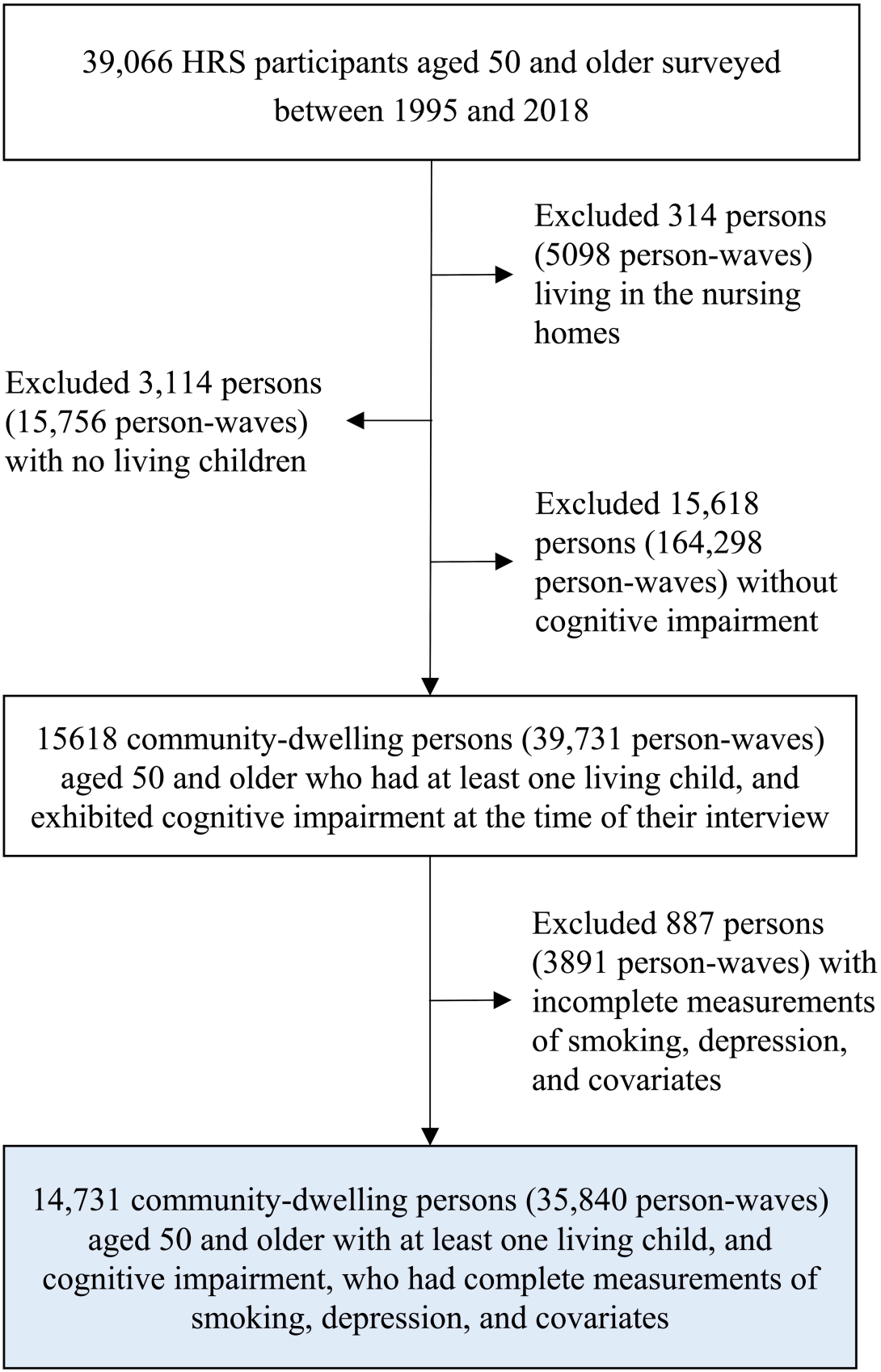
Flow chart of sample selection process *Notes*: HRS=Health and Retirement Study

Participants’ residential proximity to their children was assessed by their residential distances. In each wave, participants reported whether any children co-resided with them. For children living separately, participants specified if they lived within a 10-mile radius. Based on this data, participants fell into one of three categories: those with at least one co-residing child, those without co-residing children but with at least one child within 10 miles, and those whose children all residing more than 10 miles away. Simultaneously, we examined participants’ spousal presence by determining if they were married or partnered with a spouse present. By synthesizing these two pieces of information, we constructed our primary variable illustrating family dynamics and structure. This variable consisted of six categories representing varying levels of family support access: (1) with a spouse and co-resident children (the reference group); (2) with a spouse and children living within 10 miles; (3) with a spouse and children living beyond 10 miles; (4) without a spouse but having children co-residing; (5) without a spouse but having children living within 10 miles; and (6) without a spouse and all children living beyond 10 miles.

As for modifiable risk factors, the *Lancet* Commission Report on dementia prevention provided an established framework based on existing literature. The report estimated and ranked the population attributable fraction (PAF) of potentially modifiable risk factors for dementia using the same model, and pinpointed three modifiable risk factors for dementia that exert the most substantial influence (i.e., PAF) on dementia in later life: smoking, depressive symptoms, and social isolation. In this study, the three factors were considered as the primary outcomes. Participants who currently smoked were categorized under “smoking”. The 8-item Center for Epidemiological Studies-Depression scale (CES-D) was employed to evaluate “depressive symptoms”.^19^ Participants were asked whether they had certain feelings much of the time over the week prior to the interview (yes/no), which included 8 items such as “felt sad”, “felt lonely” and “felt depressed.” The summary score of the 8 items was constructed (range: 0-8) and a score of 3 or above was defined as having depressive symptoms.^20^ “Social isolation” was gauged using a set of six criteria that delved into participants’ social engagements with individuals, groups, and community organizations, ranging from 0 to 6.^21,22^ Drawing from an established HRS framework validated in previous literature,^21,22^ a 6-point scale was utilized to determine participants’ social isolation based on whether they (a) were unmarried; (b) lived alone; (c) had less than monthly contact with children; (d) had less than monthly contact with other family members; (e) had less than monthly contact with friends; (f) had less than monthly participation in any groups, clubs, or other social organizations. Participants providing information on at least three of these factors, but not all, were proportionally rated out of six. Those who scored above 3, falling in the top quintile, were deemed “socially isolated”.^21,22^ Data on smoking and depressive symptoms was consistently available, but social isolation metrics were limited to participants who undertook psycho-social interviews (5,951 persons and 8,376 person-waves).

To determine the association between children’s residential proximity, spousal presence and the aforementioned modifiable risk factors, we conducted mixed-effects logistic regressions. The models included individual-level random intercepts to account for the within-person correlation from multiple data points. In our analysis, we controlled for a comprehensive set of socio-demographic and health-related factors, including age, sex, race/ethnicity, education (measured in years), levels of household total wealth (categorized into quartiles), Medicare enrollment, Medicaid enrollment, Military health plan (e.g., VA) enrollment, employer-based health insurance, private health insurance, number of chronic diseases, number of children, functional limitations, and cognitive function (measured by the 27-point cognitive scale). Robust standard errors were estimated with clusters defined at the individual level.

A series of sensitivity analyses were conducted to assess the robustness of the results. First, to mitigate potential recall bias, we restricted our sample to include only participants with mild cognitive impairment, excluding those with dementia. Second, recognizing that the measure of social isolation may to some extent overlap with the concept of family support, we reanalyzed the data using two alternative measures, i.e., the subjective feelings of social isolation from others, and self-reported loneliness.^21,22^ These measures are often considered psychological manifestations of social isolation; and the analysis would help to corroborate the reliability of our findings. Lastly, we additionally adjusted for the baseline level of outcomes in the regression models to gain further insights into the directionality of the observed associations.

All analytical processes were executed in Stata 17.0, utilizing two-sided tests, with a 5% threshold designated for statistical significance.

## 3. Results

Table 1 presents the sample characteristics. On average, participants were 71.8 years old with a standard deviation of 11.1 years. Of the total, 20,868 (58.2%) were female and 18,195 (50.8%) identified as non-Hispanic White. The median number of children per participant was 3, and the median score on the 27-point cognitive scale was 9 points. Examining modifiable risk factors, 5,545 (15.5%) participants were current smokers, 12,449 (34.7%) exhibited signs of depressive symptoms, and 1,119 (13.4%) were deemed socially isolated. Participants paired with a spouse and with children residing further away generally showcased better socioeconomic status and health metrics. They typically had a higher educational attainment, greater wealth level, and fewer functional limitations than their counterparts. Conversely, those without a spouse and whose children lived far away experienced a higher prevalence of social isolation. Additionally, the prevalence of depressive symptoms was more pronounced among participants without a spouse. The differences in smoking were less discernable among these groups.

**Table 1.**
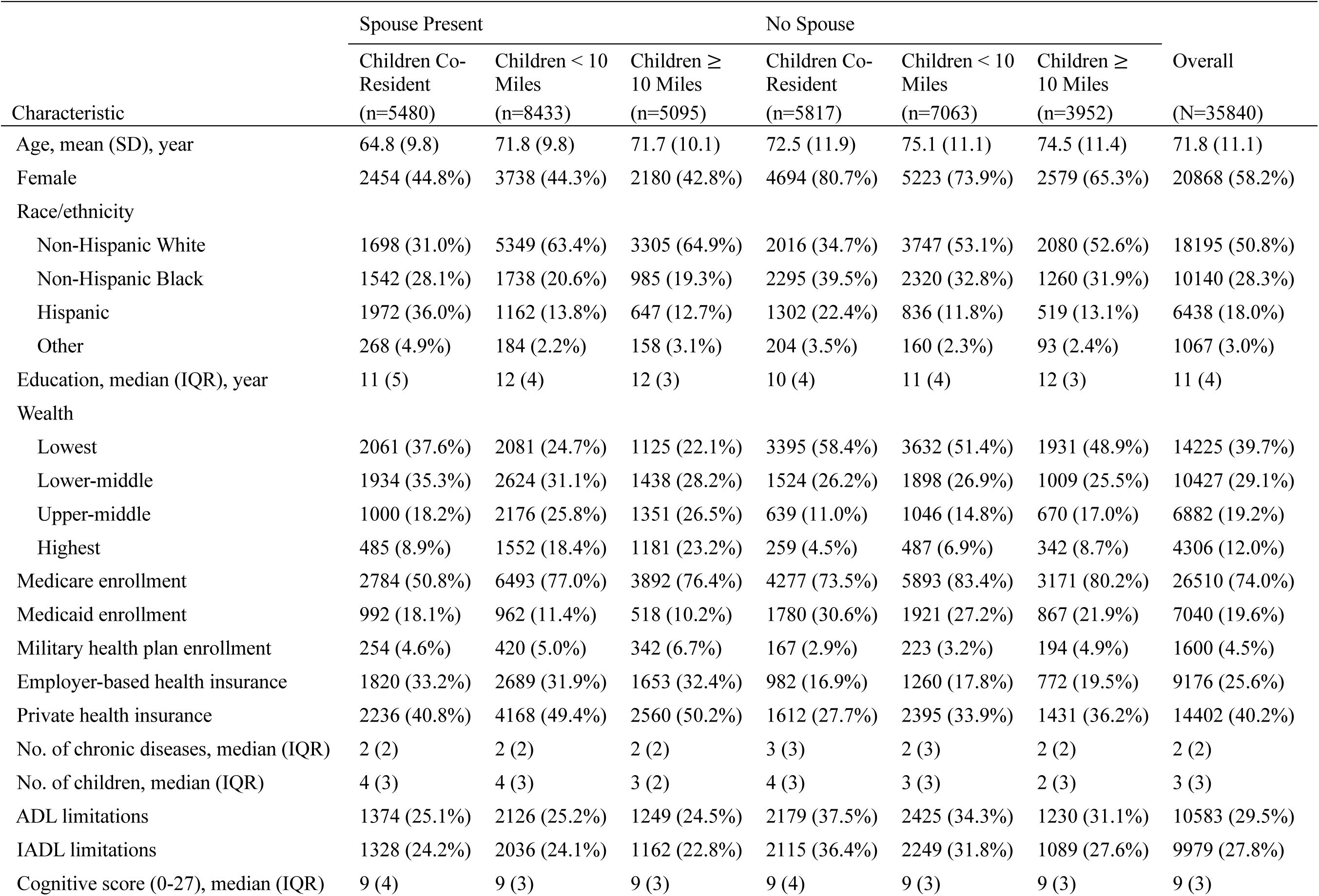
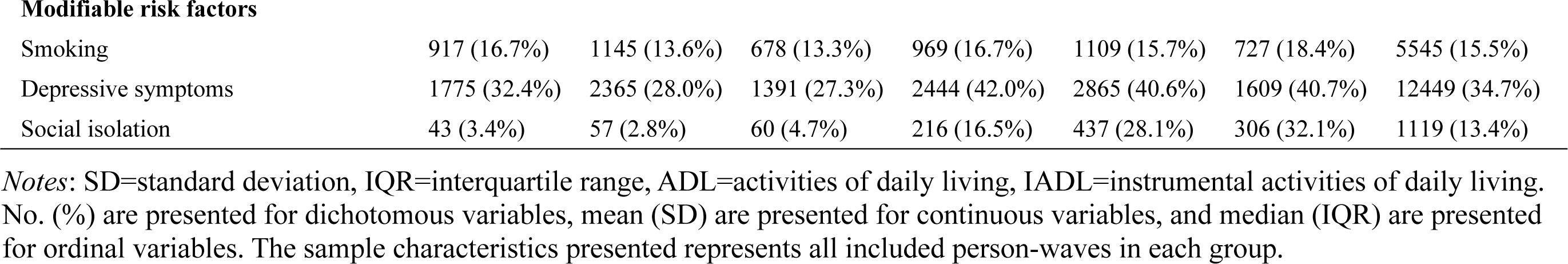
Sample characteristics by access to family support and overall, No. (%)

| Characteristic | Spouse Present |  |  | No Spouse |  |  | Overall<br>(N=35840) |
| --- | --- | --- | --- | --- | --- | --- | --- |
|  | Children Co-Resident<br>(n=5480) | Children < 10 Miles<br>(n=8433) | Children ≥ 10 Miles<br>(n=5095) | Children Co-Resident<br>(n=5817) | Children < 10 Miles<br>(n=7063) | Children ≥ 10 Miles<br>(n=3952) |  |
| Age, mean (SD), year | 64.8 (9.8) | 71.8 (9.8) | 71.7 (10.1) | 72.5 (11.9) | 75.1 (11.1) | 74.5 (11.4) | 71.8 (11.1) |
| Female | 2454 (44.8%) | 3738 (44.3%) | 2180 (42.8%) | 4694 (80.7%) | 5223 (73.9%) | 2579 (65.3%) | 20868 (58.2%) |
| Race/ethnicity |  |  |  |  |  |  |  |
| Non-Hispanic White | 1698 (31.0%) | 5349 (63.4%) | 3305 (64.9%) | 2016 (34.7%) | 3747 (53.1%) | 2080 (52.6%) | 18195 (50.8%) |
| Non-Hispanic Black | 1542 (28.1%) | 1738 (20.6%) | 985 (19.3%) | 2295 (39.5%) | 2320 (32.8%) | 1260 (31.9%) | 10140 (28.3%) |
| Hispanic | 1972 (36.0%) | 1162 (13.8%) | 647 (12.7%) | 1302 (22.4%) | 836 (11.8%) | 519 (13.1%) | 6438 (18.0%) |
| Other | 268 (4.9%) | 184 (2.2%) | 158 (3.1%) | 204 (3.5%) | 160 (2.3%) | 93 (2.4%) | 1067 (3.0%) |
| Education, median (IQR), year | 11 (5) | 12 (4) | 12 (3) | 10 (4) | 11 (4) | 12 (3) | 11 (4) |
| Wealth |  |  |  |  |  |  |  |
| Lowest | 2061 (37.6%) | 2081 (24.7%) | 1125 (22.1%) | 3395 (58.4%) | 3632 (51.4%) | 1931 (48.9%) | 14225 (39.7%) |
| Lower-middle | 1934 (35.3%) | 2624 (31.1%) | 1438 (28.2%) | 1524 (26.2%) | 1898 (26.9%) | 1009 (25.5%) | 10427 (29.1%) |
| Upper-middle | 1000 (18.2%) | 2176 (25.8%) | 1351 (26.5%) | 639 (11.0%) | 1046 (14.8%) | 670 (17.0%) | 6882 (19.2%) |
| Highest | 485 (8.9%) | 1552 (18.4%) | 1181 (23.2%) | 259 (4.5%) | 487 (6.9%) | 342 (8.7%) | 4306 (12.0%) |
| Medicare enrollment | 2784 (50.8%) | 6493 (77.0%) | 3892 (76.4%) | 4277 (73.5%) | 5893 (83.4%) | 3171 (80.2%) | 26510 (74.0%) |
| Medicaid enrollment | 992 (18.1%) | 962 (11.4%) | 518 (10.2%) | 1780 (30.6%) | 1921 (27.2%) | 867 (21.9%) | 7040 (19.6%) |
| Military health plan enrollment | 254 (4.6%) | 420 (5.0%) | 342 (6.7%) | 167 (2.9%) | 223 (3.2%) | 194 (4.9%) | 1600 (4.5%) |
| Employer-based health insurance | 1820 (33.2%) | 2689 (31.9%) | 1653 (32.4%) | 982 (16.9%) | 1260 (17.8%) | 772 (19.5%) | 9176 (25.6%) |
| Private health insurance | 2236 (40.8%) | 4168 (49.4%) | 2560 (50.2%) | 1612 (27.7%) | 2395 (33.9%) | 1431 (36.2%) | 14402 (40.2%) |
| No. of chronic diseases, median (IQR) | 2 (2) | 2 (2) | 2 (2) | 3 (3) | 2 (3) | 2 (2) | 2 (2) |
| No. of children, median (IQR) | 4 (3) | 4 (3) | 3 (2) | 4 (3) | 3 (3) | 2 (3) | 3 (3) |
| ADL limitations | 1374 (25.1%) | 2126 (25.2%) | 1249 (24.5%) | 2179 (37.5%) | 2425 (34.3%) | 1230 (31.1%) | 10583 (29.5%) |
| IADL limitations | 1328 (24.2%) | 2036 (24.1%) | 1162 (22.8%) | 2115 (36.4%) | 2249 (31.8%) | 1089 (27.6%) | 9979 (27.8%) |
| Cognitive score (0-27), median (IQR) | 9 (4) | 9 (3) | 9 (3) | 9 (4) | 9 (3) | 9 (3) | 9 (3) |
| **Modifiable risk factors** |  |  |  |  |  |  |  |
| Smoking | 917 (16.7%) | 1145 (13.6%) | 678 (13.3%) | 969 (16.7%) | 1109 (15.7%) | 727 (18.4%) | 5545 (15.5%) |
| Depressive symptoms | 1775 (32.4%) | 2365 (28.0%) | 1391 (27.3%) | 2444 (42.0%) | 2865 (40.6%) | 1609 (40.7%) | 12449 (34.7%) |
| Social isolation | 43 (3.4%) | 57 (2.8%) | 60 (4.7%) | 216 (16.5%) | 437 (28.1%) | 306 (32.1%) | 1119 (13.4%) |

*Notes:* SD=standard deviation, IQR=interquartile range, ADL=activities of daily living, IADL=instrumental activities of daily living. No. (%) are presented for dichotomous variables, mean (SD) are presented for continuous variables, and median (IQR) are presented for ordinal variables. The sample characteristics presented represents all included person-waves in each group.

Figure 2 illustrates the association between diminished access to family support and an increased prevalence of smoking, depressive symptoms, and social isolation, after regression adjustments (also see Supplementary Table S1 for numerical estimates). Relative to participants having both a spouse and co-residing children, those with a spouse but with children residing either within 10 miles (aOR: 1.40; 95% CI, 1.07-1.82) or further than 10 miles (aOR: 1.40; 95% CI, 1.04-1.90) exhibited significantly elevated odds of smoking. The absence of a spouse exacerbated these associations, and the farther away their children resided, the higher the chances of experiencing smoking, depressive symptoms, and social isolation.

**Figure 2.**
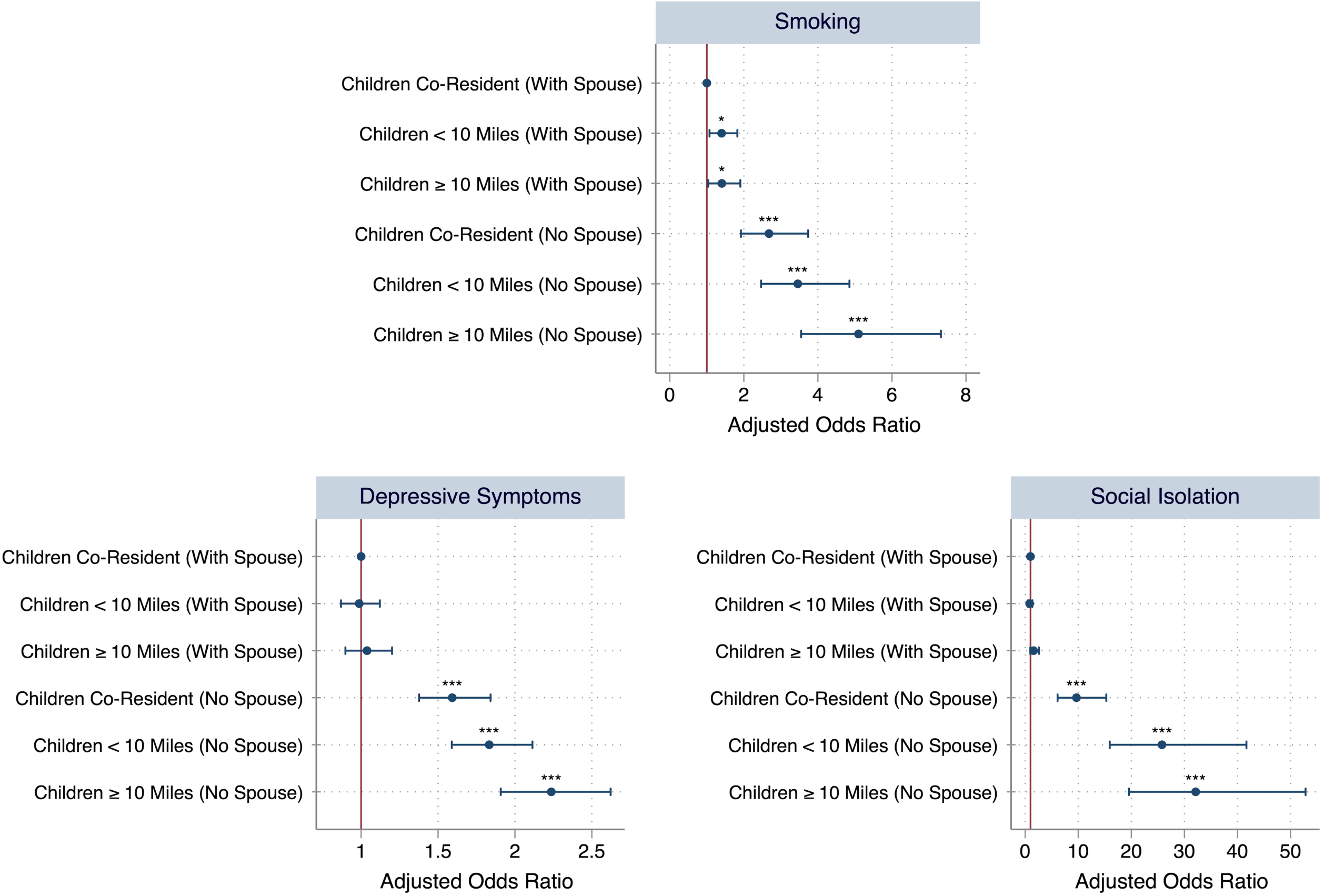
Association between family support and modifiable risk factors for older adults with cognitive impairment *Notes*: “Children Co-Resident” = any children co-resident with the respondents; “Children < 10 Miles” = any children living within 10 miles from the respondents; “Children ≥ 10 Miles” = all children living 10 miles away from the respondents. The effects were estimated using mixed effects logistic regressions, controlling for age, sex, race/ethnicity, education, wealth, Medicare enrollment, Medicaid enrollment, military health plan enrollment, employer-based health insurance, private health insurance, number of chronic diseases, number of children, ADL limitations, IADL limitations, and cognitive function. Individual-level random intercepts were included to account for within-individual correlation of multiple measurements. Robust standard errors were estimated with clusters defined at the individual level. The numerical estimates are provided in Supplementary Table S1. Asterisks denote the statistical significance of the association: *** 𝑃<0.001, ** 𝑃<0.01, * 𝑃<0.05.

Compared to the reference group (i.e., spouses were present and children co-resided), those without a spouse but having co-residing children had increased odds of smoking (aOR: 2.68; 95% CI, 1.92-3.73), depressive symptoms (aOR: 1.59; 95% CI, 1.38-1.84), and social isolation (aOR: 9.66; 95% CI, 6.11-15.28). This trend was even more pronounced for participants without a spouse and children living within a 10-mile radius, as evidenced by the heightened likelihood of smoking (aOR: 3.46; 95% CI, 2.47-4.85), depressive symptoms (aOR: 1.83; 95% CI, 1.59-2.11), and social isolation (aOR: 25.78; 95% CI, 15.94-41.69). The strongest associations were observed in those without a spouse and all children residing beyond 10 miles, with the highest odds of smoking (aOR: 5.10; 95% CI, 3.55-7.32), depressive symptoms (aOR: 2.24; 95% CI, 1.91-2.62), and social isolation (aOR: 32.13; 95% CI, 19.54-52.83). The associations between family support access and these risk factors showed a consistent gradient across all outcomes.

The gradient relationship was further corroborated through our sensitivity analyses. Specifically, our findings remained robust when we limited the sample to those without dementia (Supplementary Table S2), when we considered alternative measures of social isolation (Supplementary Table S3), and when we controlled for the baseline outcome, i.e., the baseline level of modifiable risk factors, which to some extent supports the directionality of family support reducing risk factors (Supplementary Table S4).

## 4. Discussion

The present study highlights strong associations between family support access and modifiable risk factors for dementia among older adults with cognitive impairment. By examining the proximity of children and spousal presence as indicators of family support, we reveal a notable gradient in older persons’ prevalence of smoking, depressive symptoms, and social isolation based on the accessibility of their family support.

At the core of our findings is a pronounced gradient relationship: the diminished access to family support is associated with an elevated likelihood of these risk factors. Older adults with cognitive impairment, already navigating the complexities of their condition, appear to particularly benefit from close familial bonds. This is manifested most evidently in those having both a spouse and co-residing children, who displayed the least likelihood of engaging in smoking, experiencing depressive symptoms, or being socially isolated. This underscores the invaluable protective layer that immediate family can provide, particularly in the context of cognitive challenges.

While spousal presence emerged as a substantial protective factor, the residential proximity of children introduced an additional layer of nuance. Even for participants with a spouse, a further residential distance from their children was associated with a heightened prevalence of smoking. This amplifies the significance of both spousal and child-based support in the well-being of cognitively impaired older adults.

Importantly, this study stresses the elevated vulnerability for older adults without a spouse, particularly when their children live farther away. This group exhibited the highest odds for all three risk factors, with the risks magnifying with increased distance from their children. Since only about one-fifth of older adults living alone with cognitive impairment are covered by Medicaid, this population group is largely ineligible for consistent access to publicly subsidized essential health care and social services.^4^ All these facts suggest that enhancing family or social support mechanisms for these individuals might be paramount, also a priority of the National Alzheimer’s Project Act that requires the U.S. Department of Health and Human Services (DHHS) to provide adequate supports to people with cognitive impairment.^23^ Countries have also introduced a set of direct and indirect measures to encourage adult children to live closer to older parents. In Singapore, both a Proximity Housing Grant and the Married Child Priority Scheme set aside housing subsidies for children who want to reside with or near their parents.^24,25^ In America, European countries etc., paid family leave policies aim to reduce the price of adult child-to-parent caregiving, fostering more informal care with proximity.^26^

A potential limitation of our study is its correlational nature. While the associations between family support and risk factors among cognitively impaired older adults are evident, establishing causality and illuminating the underlying mechanisms remains a challenge. We did however find from a sensitivity analysis that lower family support predicts future risk factors even when we adjusted for x and y at baseline, which suggests that the family support drives the relationship. Future research could further explore this. Interventions and policy changes, for instance, could be utilized to examine how changes in family support would affect these risk factors. Moreover, although our study contributes by examining the residential proximity of children, we did not differentiate the social ties and closeness within families, which warrants further investigation. Lastly, even though we demonstrated the robustness of our findings by using a sample with minimal cognitive impairment, recall bias and mortality bias may still exist in the analysis of cognitively impaired older adults. The significant pattern of our results remained, however, when we conducted a sensitivity analysis that excluded those with greater cognitive impairment and focused on those who were most likely to have accurate recall.

In conclusion, this study accentuates the significance of family support and their potential impact on key modifiable risk factors for dementia in older adults with cognitive impairment. Addressing these risk factors, especially in those with limited family support, can play a pivotal role in enhancing their health and well-being.

## Data Availability

All data produced in the present study are available upon reasonable request to the authors.

## Acknowledgements

This study was funded by research grants R01AG077529, R01AG067533 and U01AG032284 from the U.S. National Institute on Aging; career development award K01AG053408 from the U.S. National Institute on Aging; and grant P30AG021342 from the U.S. National Institute on Aging to the Yale Claude D. Pepper Older Americans Independence Center. The funders had no role in the study design; data collection, analysis, or interpretation; in the writing of the report; or in the decision to submit the article for publication.

---

**Supplementary Table S1.**
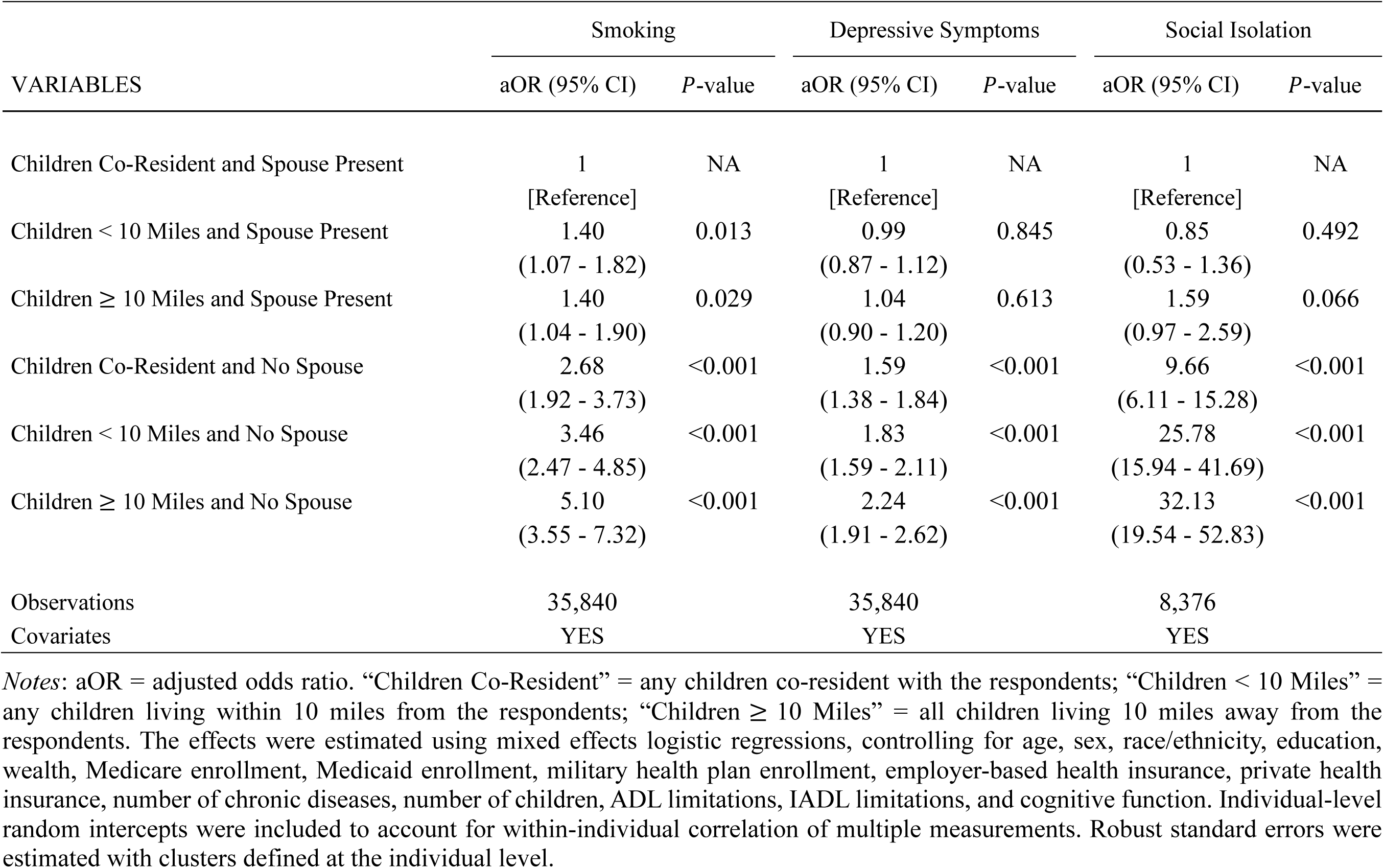
**Numerical estimates for the effects of family support on modifiable risk factors for older adults with cognitive impairment**

| VARIABLES | Smoking |  | Depressive Symptoms |  | Social Isolation |  |
| --- | --- | --- | --- | --- | --- | --- |
|  | aOR (95% CI) | P-value | aOR (95% CI) | P-value | aOR (95% CI) | P-value |
| Children Co-Resident and Spouse Present | 1<br>[Reference] | NA | 1<br>[Reference] | NA | 1<br>[Reference] | NA |
| Children < 10 Miles and Spouse Present | 1.40<br>(1.07 - 1.82) | 0.013 | 0.99<br>(0.87 - 1.12) | 0.845 | 0.85<br>(0.53 - 1.36) | 0.492 |
| Children ≥ 10 Miles and Spouse Present | 1.40<br>(1.04 - 1.90) | 0.029 | 1.04<br>(0.90 - 1.20) | 0.613 | 1.59<br>(0.97 - 2.59) | 0.066 |
| Children Co-Resident and No Spouse | 2.68<br>(1.92 - 3.73) | <0.001 | 1.59<br>(1.38 - 1.84) | <0.001 | 9.66<br>(6.11 - 15.28) | <0.001 |
| Children < 10 Miles and No Spouse | 3.46<br>(2.47 - 4.85) | <0.001 | 1.83<br>(1.59 - 2.11) | <0.001 | 25.78<br>(15.94 - 41.69) | <0.001 |
| Children ≥ 10 Miles and No Spouse | 5.10<br>(3.55 - 7.32) | <0.001 | 2.24<br>(1.91 - 2.62) | <0.001 | 32.13<br>(19.54 - 52.83) | <0.001 |
| Observations | 35,840 |  | 35,840 |  | 8,376 |  |
| Covariates | YES |  | YES |  | YES |  |

*Notes:* aOR = adjusted odds ratio. “Children Co-Resident” = any children co-resident with the respondents; “Children < 10 Miles” = any children living within 10 miles from the respondents; “Children ≥ 10 Miles” = all children living 10 miles away from the respondents. The effects were estimated using mixed effects logistic regressions, controlling for age, sex, race/ethnicity, education, wealth, Medicare enrollment, Medicaid enrollment, military health plan enrollment, employer-based health insurance, private health insurance, number of chronic diseases, number of children, ADL limitations, IADL limitations, and cognitive function. Individual-level random intercepts were included to account for within-individual correlation of multiple measurements. Robust standard errors were estimated with clusters defined at the individual level.

**Supplementary Table S2.** **Sensitivity analysis: robustness to the restriction of sample to participants with mild cognitive impairment**

| VARIABLES | Smoking |  | Depressive Symptoms |  | Social Isolation |  |
| --- | --- | --- | --- | --- | --- | --- |
|  | aOR (95% CI) | P-value | aOR (95% CI) | P-value | aOR (95% CI) | P-value |
| Children Co-Resident and Spouse Present | 1<br>[Reference] | NA | 1<br>[Reference] | NA | 1<br>[Reference] | NA |
| Children < 10 Miles and Spouse Present | 1.49<br>(1.12 - 1.99) | 0.006 | 0.97<br>(0.85 - 1.11) | 0.677 | 0.81<br>(0.48 - 1.35) | 0.411 |
| Children ≥ 10 Miles and Spouse Present | 1.50<br>(1.07 - 2.09) | 0.018 | 1.03<br>(0.88 - 1.20) | 0.722 | 1.59<br>(0.94 - 2.69) | 0.083 |
| Children Co-Resident and No Spouse | 3.28<br>(2.31 - 4.65) | <0.001 | 1.66<br>(1.41 - 1.94) | <0.001 | 10.33<br>(6.21 - 17.18) | <0.001 |
| Children < 10 Miles and No Spouse | 4.08<br>(2.84 - 5.86) | <0.001 | 1.87<br>(1.60 - 2.18) | <0.001 | 22.00<br>(12.94 - 37.40) | <0.001 |
| Children ≥ 10 Miles and No Spouse | 5.55<br>(3.77 - 8.16) | <0.001 | 2.33<br>(1.96 - 2.76) | <0.001 | 29.69<br>(17.02 - 51.78) | <0.001 |
| Observations | 28,845 |  | 28,845 |  | 6,969 |  |
| Covariates | YES |  | YES |  | YES |  |

**Supplementary Table S3.** **Sensitivity analysis: robustness to the alternative measures of social isolation**

| VARIABLES | Feel Socially Isolated |  | Loneliness |  |
| --- | --- | --- | --- | --- |
|  | aOR (95% CI) | P-value | aOR (95% CI) | P-value |
| Children Co-Resident and Spouse Present | 1<br>[Reference] | NA | 1<br>[Reference] | NA |
| Children < 10 Miles and Spouse Present | 1.17<br>(0.92 - 1.50) | 0.209 | 1.17<br>(0.83 - 1.65) | 0.382 |
| Children ≥ 10 Miles and Spouse Present | 1.50<br>(1.15 - 1.95) | 0.003 | 1.68<br>(1.16 - 2.43) | 0.006 |
| Children Co-Resident and No Spouse | 2.06<br>(1.55 - 2.74) | <0.001 | 3.31<br>(2.28 - 4.81) | <0.001 |
| Children < 10 Miles and No Spouse | 2.00<br>(1.52 - 2.64) | <0.001 | 2.91<br>(2.01 - 4.21) | <0.001 |
| Children ≥ 10 Miles and No Spouse | 1.90<br>(1.40 - 2.56) | <0.001 | 3.26<br>(2.19 - 4.84) | <0.001 |
| Observations | 6,682 |  | 6,757 |  |
| Covariates | YES |  | YES |  |

*Notes:* aOR = adjusted odds ratio. “Children Co-Resident” = any children co-resident with the respondents; “Children < 10 Miles” = any children living within 10 miles from the respondents; “Children ≥ 10 Miles” = all children living 10 miles away from the respondents. Individuals were defined as feeling socially isolated if they self-reported to feel isolated from others for some of the time or often. In addition, loneliness score (ranges 3-9) was assigned to each individual based on how frequently he/she felt 1) lacking companionship, 2) left out, and 3) isolated from others (each item ranges from 1-3: hardly ever or never=1, some of the time=2, often=3). Those in the top quintile of loneliness scores are classified as lonely. The effects were estimated using mixed effects logistic regressions, controlling for age, sex, race/ethnicity, education, wealth, Medicare enrollment, Medicaid enrollment, military health plan enrollment, employer-based health insurance, private health insurance, number of chronic diseases, number of children, ADL limitations, IADL limitations, and cognitive function. Individual-level random intercepts were included to account for within-individual correlation of multiple measurements. Sample were restricted to participants with mild cognitive impairment to reduce recall bias. Robust standard errors were estimated with clusters defined at the individual level.

**Supplementary Table S4.**
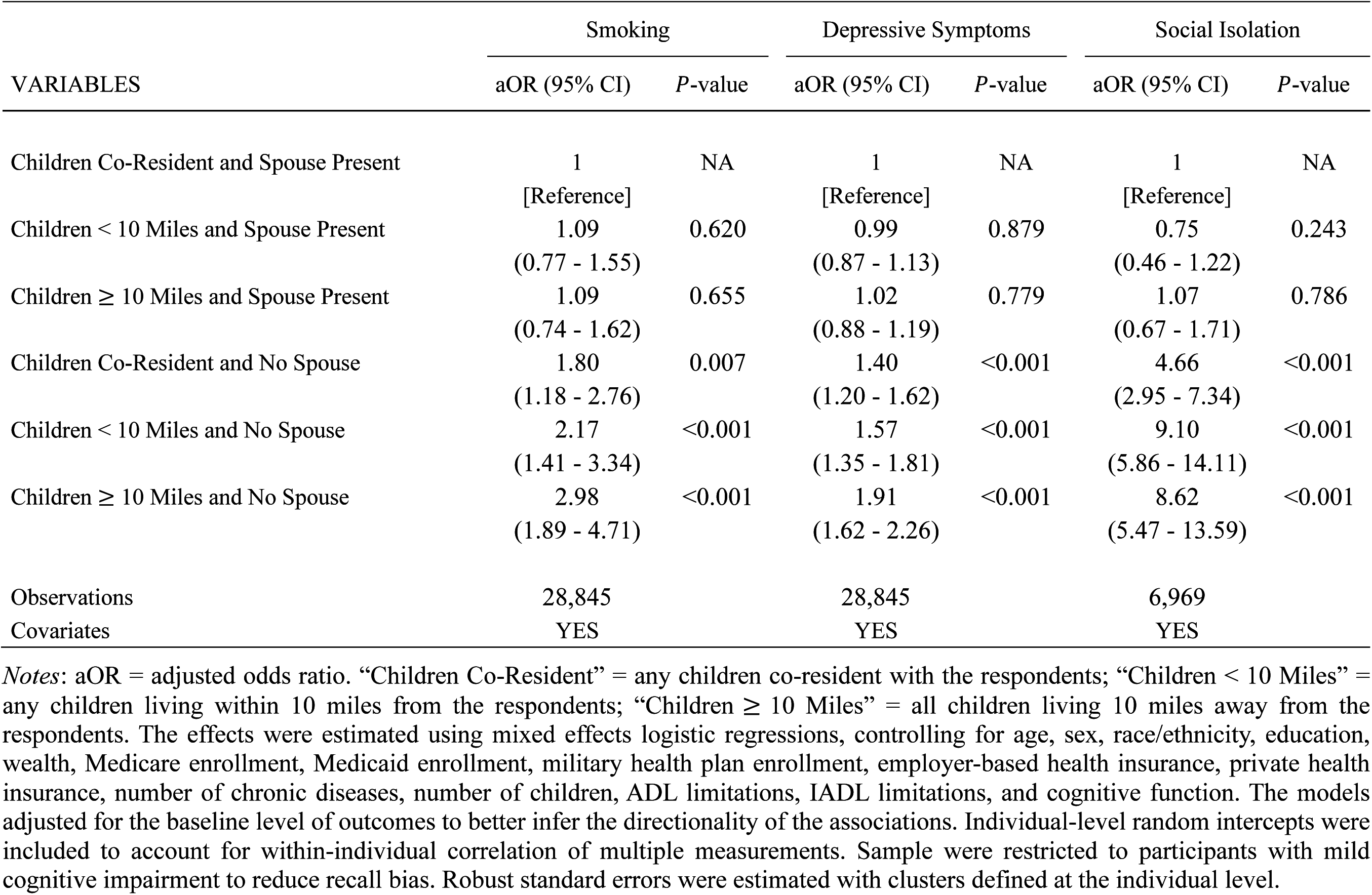
**Sensitivity analysis: robustness to the control of baseline outcomes**

